# Predictors of brain iron deposition in dementia and Parkinson’s disease-associated subcortical regions: genetic and observational analysis in UK Biobank

**DOI:** 10.1101/2025.02.11.25321877

**Authors:** Francesco Casanova, Qu Tian, Daniel S Williamson, Mitchell R Lucas, David Zweibaum, Jun Ding, Janice L Atkins, David Melzer, Luigi Ferrucci, Luke C Pilling

**Author notes:** Corresponding author: Dr Luke Pilling, College House (1.09), St. Luke’s campus, University of Exeter, Exeter, Devon, EX1 2LU, UK.

## Abstract

**Background:** Brain iron in specific subcortical regions increases risk of dementia and Parkinson’s Disease (PD). Genetic and environmental factors affect iron deposition, but the underlying mechanisms are unclear.

**Objective:** Identify risk factors and diseases associated with brain iron, and assess causality using genetics.

**Methods:** 41,581 UK Biobank participants had MRI-estimated brain iron in four dementia or PD-associated subcortical regions (Caudate, Putamen, Substantia Nigra, Thalamus). We investigated common risk factors (including adiposity, blood pressure, health behaviours, and inflammation) and diseases observationally, using covariate-adjusted regression models, and genetically, with Mendelian randomization.

**Results:** Participants diagnosed with Alzheimer’s disease, PD, or other diseases had higher brain iron. Anaemia, osteoporosis, and hyperparathyroidism were associated with lower brain iron. Higher BMI and blood pressure, history of smoking, and self-reported meat consumption, increased brain iron. Haematological parameters, inflammatory and kidney biomarkers, and calcium, were also associated.

Genetics support causal effects of depression, type-2 diabetes, and 7 other diseases with increased iron, but not Alzheimer’s disease. Evidence supports a causal effect of osteoporosis on lower iron in the substantia nigra. We found causal associations between adiposity and proteins (such as IL-6 receptor and transferrin receptor) on subcortical brain iron.

**Conclusions:** We identified causal effects for liability to type-2 diabetes, depression, and other conditions, on subcortical iron deposition, but not to Alzheimer’s disease, supportive of dementia as a consequence of brain iron deposition, not a cause. The role of adiposity reducing interventions on brain iron should be investigated. Relationships between brain iron, osteoporosis, calcium, and hyperparathyroidism warrant further investigation.

## 1 Introduction

Brain iron deposition is thought to increase risk of neurodegenerative disease,^1^ yet mechanisms are complex, with interactions between chronic diseases, hepcidin production, iron retention by macrophages, and iron absorption in the gut.^2^ We previously reported that higher plasma iron and iron deposition in specific subcortical brain regions are causally associated with increased risk of non-Alzheimer’s dementia and Parkinson’s disease.^3,4^ Brain iron deposition is present in Alzheimer’s disease and is associated with cognition, but may not be causal, with as yet no conclusive evidence supporting intervention with iron chelators.^5^ The underlying risk factors that increase brain iron deposition are not fully understood, with uncertainty over the importance of reported factors in the causal chain leading to disease.

Brain iron naturally accumulates over the lifecourse.^1^ The role of genetics in brain iron accumulation is supported by genome-wide association studies (GWAS)^6^ and studies of the iron-overload disease haemochromatosis.^7^ Type-2 diabetes and glycaemic traits increase brain iron,^8–10^ as well as BMI^11^ and cardiovascular risk factors,^12^ implicating metabolic dysfunction as a driver of iron overload. Modifiable risk factors such as smoking^8,11^ and blood pressure^13^ highlight vascular mechanisms. Evidence for environmental and behavioural factors are not consistent across studies and their causal role remains unclear.^6^

The importance of establishing causality is shown by alcohol consumption: observationally, higher consumption is associated with higher brain iron, but Mendelian Randomization (MR) analysis does not consistently support a causal effect.^14^ MR uses genetic variants as instrumental variables (proxies) for an exposure, reducing bias from reverse causation and unmeasured confounders.^15^

Our recent MR analysis supports a causal effect of iron in the thalamus on non-Alzheimer’s dementia risk, and iron in the substantia nigra, caudate and putamen on PD risk.^3,4^ In this study we aimed to identify factors causally associated with brain iron deposition in these 4 regions using observational and genetic analyses.

## 2 Methods

### 2.1 Study sample

UK Biobank (UKB) is a cohort study of ∼502,000 UK adults aged 40-70 years at baseline assessment (2006-2010). Data includes characteristics, biomarkers, genetics, and medical records. Data are available following application (www.ukbiobank.ac.uk). The Northwest Multi-Center Research Ethics Committee approved the collection and use of UKB data (Research Ethics Committee reference 11/NW/0382). Participants gave informed consent for the use of their data for health-related research purposes. Access to UKB was granted under Application Number 83534.

#### 2.1.1 MRI iron estimation

MRI data was processed centrally and imaging-derived phenotypes (IDPs) made available to analysts. We used brain iron levels estimated by the QSM method^6^, available in all 4 subcortical regions of interest (caudate, putamen, substantia nigra, and thalamus, see Supplementary Information for data fields). Mean values for right and left hemisphere measures were used. Data was available in 41,581 participants (November-2024).

#### 2.1.2 Participant characteristics, risk factors, and long-term conditions

Phenotypes from MRI assessment were age (UKB field=53), sex (field=31), assessment centre (field=54), education/qualifications (field=6138), smoking status (field=20116), alcohol intake frequency (field=1558), and “number of days per week of moderate physical activity 10+ minutes” (field=884). Some groups were combined, e.g., “secondary school” (CSEs+GCSEs) qualifications, and “4+ days per week” for moderate activity. We created a combined “number of days per week of red or processed meat consumption” phenotype using four food frequency questionnaire fields: lamb/mutton (field=1379), pork (1389), beef (1369), and processed meat (1349).

Baseline fields used were “self-reported ethnic background” (field=21000, with subgroups combined into “White,” “Asian,” “Black,” “Mixed,” and “Other” groups), and biochemistry (n=30 fields from UKB category=17518) and haematology (n=31 fields from category=100081) fields.

We ascertained the date of first diagnosis of 88 long-term conditions from hospital episode statistics (HES, censored 30-October-2022), death certificates (censored 30-November-2022), cancer registry (censored 31-December-2020), and primary care (available in ∼45% of the cohort, censored 31-May-2016 – provider dependent). The conditions comprised 83 from the GEMINI analysis of common long-term conditions^16^ plus haemochromatosis and 4 dementia phenotype (all-cause dementia, Alzheimer’s disease, non-Alzheimer’s dementia, and vascular dementia). See Supplementary Table (ST) 1 for diagnostic codes and R package ‘ukbrapR’ v0.2.8 for code to ascertain diagnoses in the UKB Research Analysis Platform (https://github.com/lcpilling/ukbrapR/).

#### 2.1.3 Olink proteomics

Plasma proteins were measured by the Pharma Proteomics Project (UKB-PPP) using an antibody-based method (Olink Explore 3072 PEA) capturing 2,923 unique proteins in 54,219 participants. This included 46,595 randomly selected participants, 6,376 selected by UKB-PPP (to enrich for specific diseases), and 1,268 from the COVID-19 repeat-imaging study. Detailed methods are published.^17^ There were 5,222 participants with baseline proteins and QSM data.

#### 2.1.4 Genotype data

Participants were genotyped using two similar (>95% shared variants, n=805,426) microarray platforms: the Affymetrix Axiom UK Biobank (n=438,427 participants) and Affymetrix UKBiLEVE (n=49,950) arrays. UKB performed genotype imputation in 487,442 participants using the Haplotype Reference Consortium and UK10K reference panels (n=∼96 million variants).^18^

### 2.2 Genetic instrumental variables

For brain iron we used our published GWAS.^3^ We used published consortia meta-analysis GWAS for AD^19^ and PD^20^, and results from GEMINI for 72 long-term conditions.^16^ For biochemistry and haematology phenotypes we used the largest available GWAS.^21^ See ST2 for list.

For each protein (n=2,923) we used the strongest genetic variant mapped to the gene coding for the assayed protein (the “cis-MR” variant) reported by UKB-PPP.^17^ Of the 2,923 analysed, 2,060 had a genome wide significant (p<5*10^−8^) cis variant. We excluded 68 with low minor allele frequency (MAF<0.1%) in the MRI sample, and 42 on the X chromosome (missing in many outcome GWAS). This left 2,018 proteins for cis-MR analysis.

We used a recent GWAS of plasma transferrin saturation (TSAT),^22^ the best marker for plasma iron content, to investigate specificity.

### 2.3 Statistical analysis

R v4.2.3 was used for all analyses.

#### 2.3.1 Observational analyses

We used linear regression models throughout, unless otherwise specified. QSM brain iron estimates were inverse rank transformed to ensure Gaussian distributions and standardized units. All models were adjusted for age at MRI assessment, sex, and MRI assessment centre. Additional adjustments are detailed in the Results (e.g., age at baseline assessment for biochemistry analysis). Benjamini-Hochberg adjustment for multiple statistical testing was performed within analysis groups. Syntax is available https://github.com/AGEexeter/paper-brain_iron_causes.

We used Fisher’s Z to compare estimates between two models using the following equation:

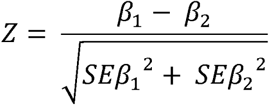

where *β*_1_ is the estimate from model 1 and *β*_2_ is the estimate from model 2, and *SEβ*_1_ is the standard error for *β*_1_ and *SEβ*_2_ is the standard error for *β*_2_.

#### 2.3.2 Mendelian randomization

For each exposure we used associated genetic variants as instrumental variables and identified associations with brain iron (QSM) using the above published GWAS. Multi-allelic SNPs were excluded. The main MR estimate is from inverse-variance weighted (IVW) analysis, which assumes there is no unbalanced horizontal pleiotropy. Additional analyses are performed to test MR assumptions, including: weighted median (assumes <50% of the weight in the analysis comes from invalid instruments); MR-Egger (assumes the genetic variants’ effect is not correlated with any pleiotropic effect on the outcome); MR-Egger (intercept term) unlike IVW the MR-Egger incept is not fixed at zero, therefore deviation from the null indicates pleiotropy. We used R packages MRlap^23^ (v0.0.3.3) and TwoSampleMR (v0.6.6) for MR analysis. MRlap performs the IVW analysis, adjusting for sample overlap and weak instrument bias (providing “corrected” betas and standard errors). TwoSampleMR performed sensitivity analyses (e.g., leave-one-out to investigate outliers). Full results available https://github.com/AGEexeter/paper-brain_iron_causes.

## 3 Results

We used data from 41,581 UK Biobank (UKB) participants with MRI-estimated brain iron (QSM) available at the time of analysis (November 2024). The mean age was 64·2 years (SD 7·74); 53% were female (Table 1).

**Table 1.** UK Biobank participant characteristics.

|  |  | **Overall**<br>**(N=41,581)** |
| --- | --- | --- |
| **Age at assessment** | Mean (SD) | 64.2 (7.74) |
|  | Median [Min, Max] | 65.0 [45.0, 83.0] |
| **Sex** | Female | 22,050 (53.0%) |
|  | Male | 19,531 (47.0%) |
| **Ethnicity** | White | 40,252 (96.8%) |
|  | Mixed | 195 (0.5%) |
|  | Asian | 541 (1.3%) |
|  | Black | 262 (0.6%) |
|  | Other | 212 (0.5%) |
|  | Missing | 119 (0.3%) |
| **Highest qualification** | Primary school | 2616 (6.3%) |
|  | Secondary school | 9561 (23.0%) |
|  | Higher qualification | 28,977 (69.7%) |
|  | Missing | 427 (1.0%) |
| **Smoking status** | Never | 25,859 (62.2%) |
|  | Previous | 13,933 (33.5%) |
|  | Current | 1357 (3.3%) |
|  | Missing | 432 (1.0%) |
| **Body mass index (BMI)** | Mean (SD) | 26.5 (4.36) |
|  | Median [Min, Max] | 25.9 [13.4, 58.0] |
|  | Missing | 1182 (2.8%) |
| **BMI (categories)** | Normal | 16,088 (38.7%) |
|  | Overweight | 16,695 (40.2%) |
|  | Obese | 7,314 (17.6%) |
|  | Underweight | 302 (0.7%) |
|  | Missing | 1,182 (2.8%) |
| **Waist circumference (cm)** | Mean (SD) | 88.3 (12.6) |
|  | Median [Min, Max] | 88.0 [53.0, 152] |
|  | Missing | 1,076 (2.6%) |
| **Waist: Hip Ratio** | Mean (SD) | 0.875 (0.0883) |
|  | Median [Min, Max] | 0.878 [0.534, 1.40] |
|  | Missing | 1,077 (2.6%) |
| **Waist: Hip Ratio**<br>**(categories)** | Normal | 30,389 (73.1%) |
|  | Overweight | 10,115 (24.3%) |
|  | Missing | 1,077 (2.6%) |
| **Systolic Blood Pressure**<br>**(mmHg)** | Mean (SD) | 139 (18.8) |
|  | Median [Min, Max] | 138 [76.5, 238] |
|  | Missing | 6,437 (15.5%) |

See Methods for details on UK Biobank fields corresponding to the above traits.

### 3.1 Risk factors

#### 3.1.1 Observational associations

Age was strongly, positive associated with QSM in the putamen and caudate (putamen standard deviation change per year [beta]=0·043: 95% Confidence Intervals 0·042 to 0·045, p=2*10^−1128^), moderately in the substantia nigra (0·002: 0·001 to 0·004, p=2*10^−4^), yet negatively in the thalamus (−0·003: −0·004 to −0·002, p=4*10^−7^). QSM values were higher in males in all 4 regions (e.g., thalamus beta=0·44: 0·42 to 0·46, p=5*10^−454^). BMI, waist circumference, and waist:hip ratio were strongly, positively associated with QSM in all 4 regions (Figure 1; ST3).

**Figure 1.**
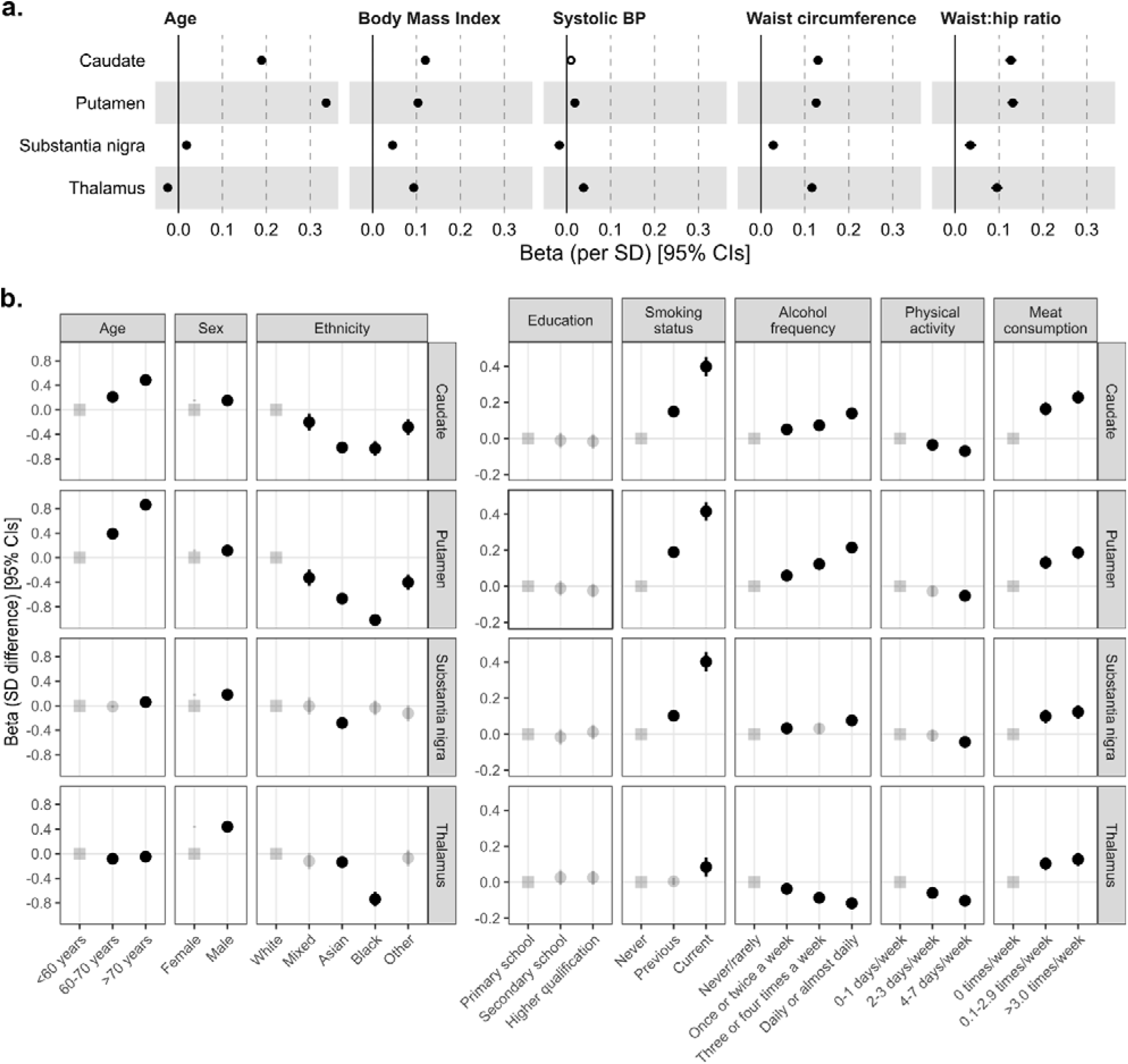
Participant characteristics at MRI assessment, associations with subcortical QSM

Self-reported participant characteristics were also strongly associated with subcortical QSM in the 4 regions (Figure 1). Self-reported ethnic background had the largest effect: black participants had >1 SD lower QSM in the putamen, compared to those self-reporting white ethnicity (beta=−1·01: −1·12 to 0·90, p=5*10^−68^). QSM values were higher in participants self-reporting higher red/processed-meat intake, and in current smokers. Higher physical activity was associated with lower QSM. Higher alcohol intake was associated with higher iron in 3 regions, yet lower in the thalamus (beta=−0·12: −0·15 to −0·08, p=3*10^−12^). All reported associations were significant after adjustment for multiple statistical testing (ST3).

From the baseline haematology panel (measured 8-14 years prior to MRI), the number of white blood cells (and subtypes) were associated with higher QSM-estimated iron in all 4 subcortical regions (Figure 2; ST4). Additionally, higher haemoglobin, mean red cell volume, and other parameters, yet lower red cell distribution width and platelet counts, were associated with higher QSM. From the baseline biochemistry panel, higher triglycerides, urate, and cystatin c were among those positively associated with QSM, whilst HDL cholesterol had the largest negative effect (Figure 2; ST4).

**Figure 2.**
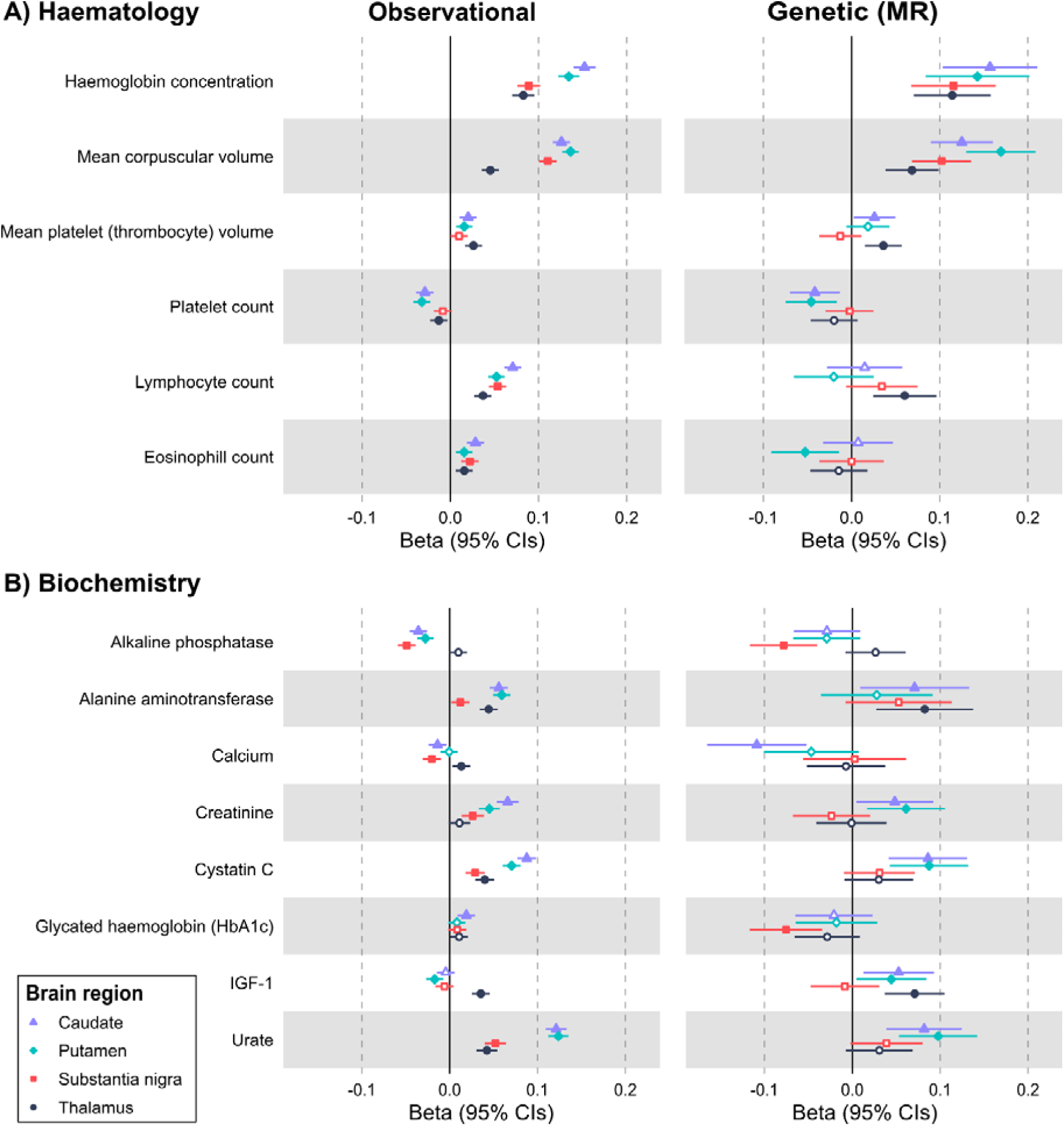
Baseline biomarkers with genetic evidence for a causal effect on QSM brain iron

Within the 2,923 proteins assayed at baseline, 107 were significantly (Bonferroni-corrected p<1·7*10^−5^) associated with at least one QSM phenotype in the 5,222 participants with MRI and proteomics data (Figure S1; ST5). Brevican core protein (BCAN) had the strongest association (beta-putamen=−0·175: −0·20 to −0·15, p=1*10^−30^). Associated proteins included those implicated in neurology (e.g., OMG), iron metabolism (TF), metabolism (LEP), and inflammation (TNFRSF10B).

Results are from linear regression models in up to 41,581 UK Biobank participants, adjusted for age at MRI assessment, sex, and MRI assessment centre. Panel A) prevalent associations between quantitative traits and QSM values (beta is the standard deviation [SD] difference in QSM per SD of the trait). Panel B) prevalent associations between categorical traits and QSM values (beta is the SD difference in QSM at MRI assessment compared to the reference category for each factor [indicated with the ‘square’]: semi-transparent points indicate the association was non-significant after adjustment for multiple testing [Benjamini-Hochberg-adjusted p>0.05]). Note axis scale differs between the panels. Lines are the 95% Confidence Intervals. See ST3 and Methods for details, including the UK Biobank fields used (some categories have been combined e.g., highest education level attained).

Observational associations are from linear regression models in up to 39,544 UK Biobank participants, adjusted for age at baseline assessment, age at MRI assessment, sex, baseline assessment centre, and MRI assessment centre. The beta is the standardized coefficient, representing the standard deviation (SD) difference in QSM at MRI assessment per standard deviation increase in baseline biomarker. Genetic associations are from Mendelian randomization analysis. The Beta is the MRlap corrected IVW estimate (i.e., the beta per SD of genetically instrumented exposure, corrected for sample overlap and weak instrument bias). Only haematology and biochemistry parameters with a significant MR effect on QSM in at least one region after Benjamini-Hochberg FDR correction are shown. See Methods and ST4 and 7 for details.

#### 3.1.2 Genetic associations

Significant observational associations were repeated using MR, where possible, to estimate causal effects.

Higher BMI was casually associated with higher brain QSM in all brain areas (Table 2; ST6). Waist circumference and waist:hip ratio were associated with higher QSM in the putamen, caudate and thalamus, but not the substantia nigra. Results do not support causal effects (FDRp>0.05) for SBP, HDL, LDL or triglycerides on QSM in these regions (ST6 and ST7). Results support causal effects for haematology and biochemistry markers (FDRp<0.05), e.g., haemoglobin and mean corpuscular volume with all regions (Figure 2; ST7). Other markers had region-specific associations: platelet count, cystatin C, urate and caudate were only associated with putamen QSM; alanine aminotransferase, IGF1, lymphocyte count, and mean platelet thrombocyte volume were only associated with thalamus QSM; alkaline phosphatase levels and hba1c were only associated with substantia nigra QSM; creatinine with putamen QSM; and calcium and IGF1 with caudate QSM (Figure 2 and ST7).

**Table 2.** Causal effects on QSM brain iron estimated by Mendelian randomization.

| **Exposure** | **Outcome region** | **Beta** | **SE** | **p-value** |
| --- | --- | --- | --- | --- |
| BMI | Caudate | 0.165 | 0.027 | 4.98E-10 |
|  | Putamen | 0.145 | 0.027 | 6.15E-08 |
|  | Substantia nigra | 0.079 | 0.028 | 4.84E-03 |
|  | Thalamus | 0.117 | 0.027 | 1.45E-05 |
| Waist circumference | Caudate | 0.177 | 0.030 | 2.38E-09 |
|  | Putamen | 0.212 | 0.031 | 1.45E-11 |
|  | Substantia nigra | 0.040 | 0.033 | 2.28E-01 |
|  | Thalamus | 0.133 | 0.030 | 7.04E-06 |
| Depression | Caudate | 0.364 | 0.117 | 1.86E-03 |
|  | Putamen | 0.258 | 0.132 | 5.15E-02 |
|  | Substantia nigra | 0.460 | 0.129 | 3.71E-04 |
|  | Thalamus | -0.070 | 0.114 | 5.41E-01 |
| Type 2 diabetes | Caudate | 0.116 | 0.030 | 9.55E-05 |
|  | Putamen | 0.093 | 0.029 | 1.42E-03 |
|  | Substantia nigra | 0.043 | 0.031 | 1.58E-01 |
|  | Thalamus | 0.060 | 0.025 | 1.84E-02 |
| Iron deficiency anaemia | Caudate | -0.358 | 0.245 | 1.43E-01 |
|  | Putamen | -0.164 | 0.332 | 6.21E-01 |
|  | Substantia nigra | -0.802 | 0.235 | 6.63E-04 |
|  | Thalamus | -0.278 | 0.239 | 2.44E-01 |
| Osteoarthritis | Caudate | 0.292 | 0.098 | 2.76E-03 |
|  | Putamen | 0.076 | 0.097 | 4.34E-01 |
|  | Substantia nigra | 0.108 | 0.109 | 3.21E-01 |
|  | Thalamus | 0.277 | 0.112 | 1.30E-02 |
| Osteoporosis | Caudate | -0.111 | 0.048 | 2.12E-02 |
|  | Putamen | -0.067 | 0.063 | 2.87E-01 |
|  | Substantia nigra | -0.257 | 0.047 | 3.74E-08 |
|  | Thalamus | 0.045 | 0.049 | 3.53E-01 |
| Psoriasis | Caudate | 0.064 | 0.014 | 5.36E-06 |
|  | Putamen | 0.056 | 0.017 | 9.03E-04 |
|  | Substantia nigra | 0.025 | 0.015 | 1.04E-01 |
|  | Thalamus | -0.045 | 0.018 | 1.37E-02 |
| Transient ischaemic attack | Caudate | 0.599 | 0.239 | 1.24E-02 |
|  | Putamen | 1.137 | 0.286 | 7.06E-05 |
|  | Substantia nigra | 0.378 | 0.238 | 1.13E-01 |
|  | Thalamus | 0.120 | 0.238 | 6.14E-01 |

Results from MRlap. The Beta, Standard Error (SE), and p-value are the estimates corrected for sample overlap and weak instrument bias. All exposures listed were significantly associated with QSM in at least one region after adjustment for multiple statistical testing (Benjamni-Hochberg $p < 0.05$ ). See Results and ST6 and ST12 for further details.

For behavioural phenotypes where a GWAS of the trait was unavailable, we used the closest phenotype where a GWAS was available (see Methods). Higher educational attainment was associated with lower caudate and putamen QSM. “Ever smoked” and higher alcohol consumption were causally associated with higher thalamus QSM, whereas overall physical activity was not associated (ST6).

Of 2,018 protein assays with a reported cis-variant^17^ 106 were significantly associated with at least one QSM trait from published GWAS^3^ after FDR correction (Benjamini-Hochberg-adjusted p<0·05) (ST8). Two proteins were associated with QSM in all four brain regions (CDSN and DBI), 16 with 3 regions, 20 with 2 regions, and 68 with one region only. Conversely, the QSM-region with most significant protein associations was the putamen (n=52), followed by the substantia nigra (n=46), caudate (n=43) and thalamus (n=23; ST8).

Using FUMA, we identified the following tissues where the above proteins with a causal effect on QSM (MRI-estimated brain iron) are differentially expressed (FDRp<0·05): adipose tissue, spleen, lung, testis and oesophagus (ST9). Additionally, gene ontology biological processes over-represented included iron metabolism, immune process and its regulation, inflammation and cytokine production, cell adhesion, and regulation of gene expression (ST10).

### 3.2 Long-term conditions

#### 3.2.1 Observational associations

We estimated associations between 88 prevalent long-term conditions (diagnoses) and QSM in the 4 subcortical regions. 48 were associated with at least one region (Figure 3 and ST11) after multiple testing correction (FDRp<0·05): conditions where QSM was higher in diagnosed individuals included Alzheimer’s disease, Parkinson’s disease, type-2 diabetes, and haemochromatosis. Four conditions were associated with lower QSM in at least one region: anaemia, osteoporosis, ulcerative colitis, and hyperparathyroidism.

**Figure 3.**
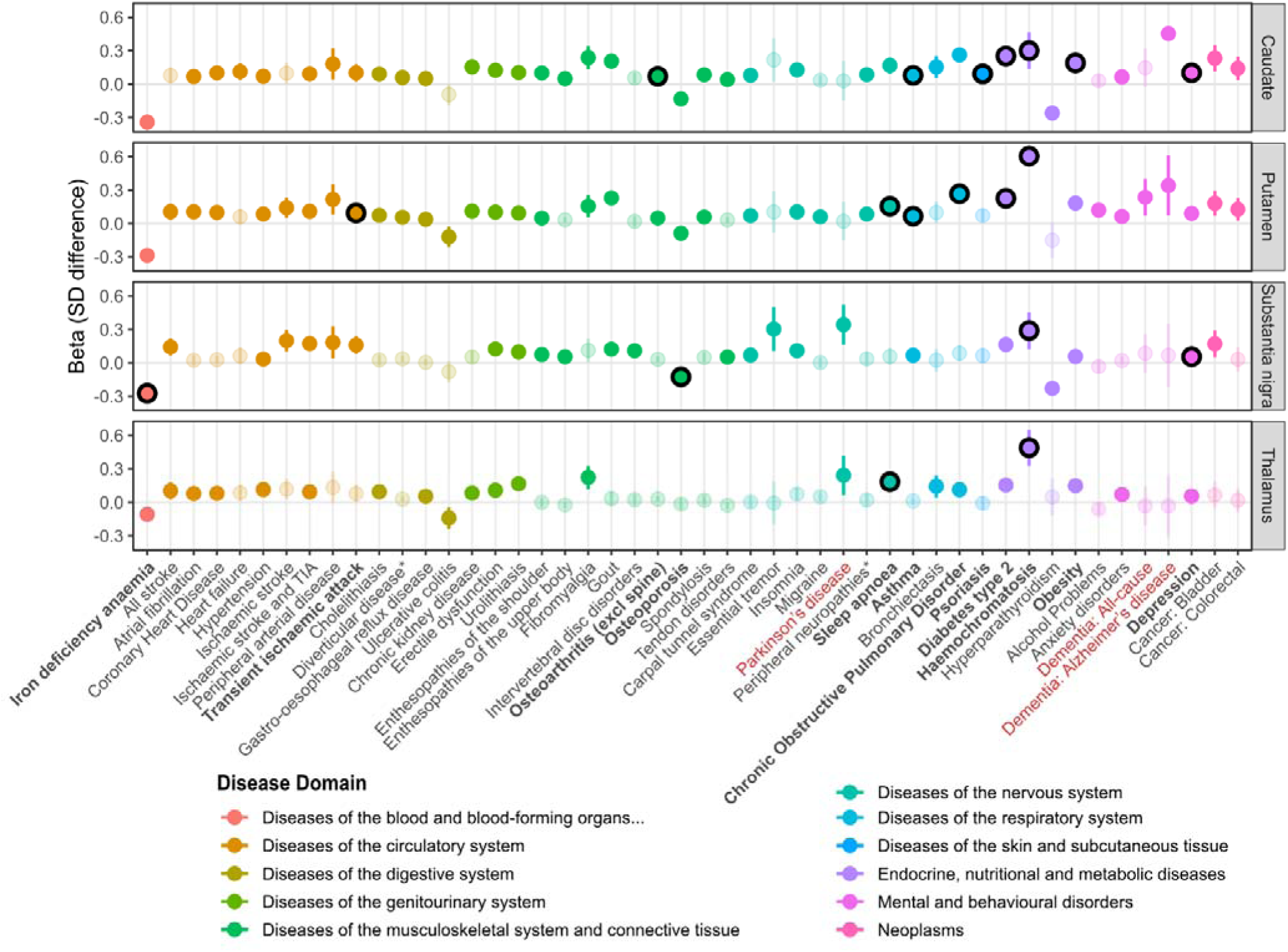
Prevalent diseases associated with subcortical QSM

Most associations remained consistent after adjustment for additional covariates (ethnic background, education [highest qualification], smoking status, alcohol [days per week], physical activity [days per week with moderate activity], meat consumption [days per week with red or processed meat intake], waist circumference, systolic blood pressure). Using Fisher’s Z to formally test for significant differences in estimates between Model 1 and Model 2, associations between diagnosed obesity and QSM were most attenuated (expected, given the adjustment for waist circumference), whilst hypertension, type-2 diabetes, gout, COPD and depression were only partially attenuated (Figure S2; ST11).

We also investigated whether prevalent disease associations differed between males and females (using Fisher’s Z): the majority of disease-QSM associations are not significantly different (Figure S3; ST11), yet some associations were stronger in females compared to males (including tendon disorders, enthesopathies, bursitis, migraine, colorectal cancer, anxiety disorders, and depression), where others were stronger in males (including schizophrenia, type-2 diabetes, hypertension, and non-melanoma skin cancer).

#### 3.2.2 Genetic associations

Mendelian Randomization of the disease-QSM associations support causal effects of genetic liability to asthma, type-2 diabetes, and psoriasis on caudate and putamen QSM (Figure 3; Table 2; ST12). Results also support causal effects of: depression on caudate and substantia nigra QSM; COPD, sleep apnoea, and TIA on putamen QSM; osteoarthritis and intervertebral disk on caudate QSM; osteoporosis, sleep apnoea, and anaemia on substantia nigra QSM; and migraine on thalamus QSM.

For haemochromatosis we investigated the known disease-causing variants *HFE* C282Y and H63D, and estimated associations with QSM, comparing levels in the YY homozygotes (n=217) to the CC/HH individuals (i.e., without either allele, n=22,961). The YY homozygotes had higher QSM in all 4 regions, with the largest effect in the putamen (beta=0·90: 0·77 to 1·02, p=2*10^−45^). Results were consistent across males and females, and in sensitivity analyses excluding those with diagnosed haemochromatosis (ST13).

Results from linear regression models in 41,581 UK Biobank participants, adjusted for age at MRI assessment, sex, and MRI assessment centre. The beta represents the standard deviation (SD) difference in QSM between the diagnosed participants and those without a diagnosis at the time of MRI assessment. Diseases are coloured according to their primary ICD-10 chapter domain. Semi-transparent points indicate the association was non-significant after adjustment for multiple testing (Benjamini-Hochberg-adjusted p>0.05). Lines are the 95% Confidence Intervals. Black outlines (and bold disease names) indicate a significant causal effect was identified in Mendelian randomization analysis (FDR p<0.05). 48 of the 88 diseases tested are shown, where there was a significant association with at least one QSM region: for further details (including full disease definitions where an abbreviated form was shown in the plot *) and results see ST11.

### 3.3 Mendelian randomization sensitivity analyses

Heterogeneity in variant-effects is common, as indicated by statistically significant Q-statistics, suggesting pleiotropic pathways (ST14). For example, some baseline haematology and biochemistry markers such as for haemoglobin and mean corpuscular volume have MR-Egger intercept p<0.05. Other results warranting cautious interpretation and further investigation include alkaline phosphatase, HbA1c, and IGF1 (ST15). Extended plots and results available https://github.com/AGEexeter/paper-brain_iron_causes.

We investigated the effect of outliers using leave-one-out analysis. For calcium there is a single larger-effect variant rs34408666 (Figure S4) yet the estimate was consistent when excluding rs34408666 (caudate-beta=−0·102, se=0·029, p=0·0005). Similarly, removing larger-effect variants for alkaline-phosphatase∼substantia-nigra (rs10916988), urate-caudate and putamen (rs4697708), asthma-caudate and putamen (rs1047989), COPD-putamen (rs11852372), diabetes-caudate and putamen (rs72826075), did not meaningfully alter results (see Figures S4 to S12). For other baseline haematology and biochemistry results there was no evidence of pleiotropy (ST15).

For long term conditions and risk factors there was no evidence that pleiotropy affected out results except for WHR on substantia nigra, depression and caudate, and osteoporosis on substantia nigra, where the MR-Egger intercept was significant (ST16). For the risk factors we only found evidence of pleiotropy for WHR-thalamus (ST17).

## 4 Discussion

We here demonstrate that adiposity (both BMI and waist circumference), causally increases MRI-estimated brain iron in subcortical regions implicated in dementia or Parkinson’s disease. We show that the pathway from adiposity to brain iron is linked to inflammation, and suggest a role for impaired glycaemic control. Our findings do not support causal effects for other cardiovascular risk factors such as high blood pressure.

Adiposity is observationally associated with brain iron in ageing mice.^11^ Visceral adiposity increased inflammation, hepcidin production, and brain iron deposition,^24^ highlighting complex interactions between hepcidin in the brain and iron, though evidence in humans is needed. Our genetic evidence supports a causal effect of adiposity (using BMI and waist circumference measures) on subcortical iron deposition. Though we lack data on hepcidin, this mechanism is supported by our report of the role of central adiposity in haemochromatosis,^25^ where HFE (homeostatic iron regulator) is responsible for detecting serum iron and regulates hepcidin.

The role of inflammation on hepcidin and iron is complex: inflammation increases hepcidin production leading to intracellular iron retention, which can cause anaemia,^2^ yet inflammation activates microglia in the brain, leading to iron deposition.^1^ A role for inflammation in brain iron deposition is supported by our proteomics results where genetically predicted higher IL6R (expressed on leukocytes and regulates the cellular response to the multifunctional cytokine IL6^26^) was associated with higher iron in the caudate. Pathway analysis also implicated the immune system and its regulation in brain iron deposition, as well as our finding that genetic liability to psoriasis and asthma increased QSM-estimated brain iron.

Consistent with previous literature linking brain iron deposition and neurodegenerative disease^1^ we observed that participants diagnosed with Alzheimer’s disease or Parkinson’s disease had higher brain iron, despite small numbers with diagnoses (n=47 and n=117, respectively). Interestingly, different regions are implicated, highlighting specific disease pathology. We did not find evidence supporting a causal effect of liability to dementia or Parkinson’s on QSM, consistent with the disease being secondary to iron deposition. We previously reported that brain iron in the 4 specific subcortical regions here studied have a causal effect on either Parkinson’s disease (caudate, putamen, and substantia nigra) or non-Alzheimer’s dementia (thalamus and putamen).^3^ Though we did not observe higher iron in the participants diagnosed with non-Alzheimer’s dementia (or vascular dementia specifically), the number are very low (n=77 and n=22, respectively) resulting in wide confidence intervals, rendering it difficult to draw firm conclusions regarding non-significant associations. Overall, our results are consistent with subcortical iron deposition occurring in participants diagnosed with dementia or Parkinson’s disease, and support a causal role of iron deposition.

Previous observational studies report associations between glycaemia and brain iron^8–10^. Our results support a causal effect of genetic liability to type-2 diabetes and iron deposition in the caudate and putamen. We speculate that microvascular dysfunction may be the mechanism, as opposed to macrovascular disease, because we found no associations between coronary heart disease or systolic blood pressure with brain iron.

Our observational results show that patients diagnosed with depression have higher brain iron, supporting previous studies implicating iron homeostasis in depression.^27^ We extend this by showing that genetic liability for depression is associated with higher brain iron, supporting a causal association. Given our above findings of a causal link between higher BMI and brain iron, and published evidence of a genetic link between BMI and depression,^28^ the relationship between depression, adiposity, and iron, warrants further investigation.

As expected, we found strong evidence that iron-related metabolism has an important role in determining brain QSM, with both observational and genetic evidence. We previously reported that the haemochromatosis-associated *HFE* C282Y variant increased brain iron estimated by the R2* method,^7^ which we here confirm with the newer QSM method.^6^ Our results highlight that a balance between high and low iron has to be achieved: we show that genetic liability for anaemia is associated with lower brain QSM, and whilst it is known that high iron is a risk factor for neurodegenerative diseases^3,4^ it has also been shown that pathologically low iron is a risk factor^29^. The association between red meat (a high-iron food) consumption and brain QSM has been previously reported^6^, which we extend by demonstrating the independent effects of red/processed meat consumption on brain iron deposition after adjustment for many other health and socio-economic factors.

Osteoporosis and hyperparathyroidism diagnoses were also associated with lower iron in the substantia nigra and caudate. Additionally, we observe that higher baseline calcium was associated with lower iron in the same regions. Calcium, like iron, has essential functions but high levels can be harmful.^30^ The relationship between calcium and iron is complex: the parathyroid glands secrete parathyroid hormone (PTH) to regulate calcium levels in the blood, and iron deposition in the glands can cause PTH dysregulation.^31^ Additionally, higher dietary calcium can inhibit non-heme iron absorption via shared transporters (e.g., DMT1).^32^ A study of 106 AD patients found higher iron in the cerebral spinal fluid yet lower calcium, possibly due to calcium reuptake by cells in response to the higher iron, leading to cell death.^33^ Our genetic analysis supports a causal effect for osteoporosis and serum calcium on lower brain iron. Statistical power for the analysis of hyperparathyroidism was limited. Our results highlight the important dynamic between iron and calcium, and the need for further work to understand the links to iron deposition in the substantia nigra and caudate.

UKB is an exceptional resource, but has limitations. Principally, the 5% response rate at baseline and known higher socio-economic demographic than the general population; yet pathological findings are generalisable to the UK population.^34^ QSM estimated brain iron from MRIs may be influenced by other factors.^6^ Diagnoses are primarily from secondary care (hospital inpatient admissions); future work integrating more primary care diagnoses are required, especially for non-hospitalising conditions. MR evidence can support causal effects of risk factors or diseases on brain iron, but relies on strong assumptions and can be biased due to pleiotropy: we investigated this in sensitivity analysis and overall the genetic and observational findings are consistent, but results should be interpreted cautiously, especially where there is evidence of heterogeneity.

To conclude, we have shown that higher adiposity and inflammation, and lower calcium, are significant causal risk factors for iron deposition in specific subcortical brain regions. Genetic liability to type-2 diabetes, depression, and other conditions, also affect subcortical iron deposition. The role of adiposity reducing interventions on brain iron should be investigated. The relationship between brain iron, osteoporosis, calcium, and hyperparathyroidism warrants further investigation.

## 5 Acknowledgements

This research has been conducted using the UK Biobank Resource, under application 83534. This work uses data provided by patients and collected by the NHS as part of their care and support, Copyright © (2024), NHS England. Re-used with the permission of the NHS England [and/or UK Biobank]. All rights reserved. The authors wish to thank the UK Biobank participants and coordinators for this unique dataset. The authors would like to acknowledge the use of the University of Exeter High-Performance Computing (HPC) facility in carrying out this work. For the purpose of open access, the author has applied a ‘Creative Commons Attribution (CC BY)’ licence to any Author Accepted Manuscript version arising from this submission.

## 6 Author Contributions

Francesco Casanova (formal analysis, investigation, writing - original draft, visualisation). Qu Tian (conceptualization, investigation, writing - original draft, funding acquisition, project administration). Daniel Williamson (methodology, writing - review & editing). Mitchell Lucas (methodology, writing - review & editing). David Zweibaum (methodology, writing - review & editing). Jun Ding (methodology, writing - review & editing). Janice Atkins (conceptualization, methodology, writing - review & editing). David Melzer (conceptualization, funding acquisition, project administration, writing - review & editing). Luigi Ferrucci (conceptualization, funding acquisition, project administration, resources, supervision, writing - original draft). Luke Pilling (conceptualization, funding acquisition, project administration, resources, supervision, writing - original draft, investigation, visualisation, formal analysis, data curation).

## 7 Statements and declarations

## 7.1 Ethical considerations

The Northwest Multi-Center Research Ethics Committee approved the collection and use of UK Biobank data (Research Ethics Committee reference 11/NW/0382).

## 7.2 Consent to participate

UK Biobank participants gave informed, written consent for the use of their data for health-related research purposes.

## 7.3 Consent for publication

Not applicable.

## 7.6 Data availability

Individual level data is available on application to UK Biobank (www.ukbiobank.ac.uk). Syntax is available https://github.com/AGEexeter/paper-brain_iron_causes.

