## Supplementary Information for "Predictors of brain iron deposition in dementia and Parkinson’s disease-associated subcortical regions: genetic and observational analysis in UK Biobank"

**Predictors and causal risk factors of MRI-estimated iron in four subcortical brain regions implicated in dementia and Parkinson’s disease**

Casanova *et al.*

**Supplementary Information**

Specific supplementary figures mentioned in the manuscript are included here. Other supplementary results files, sensitivity analyses, results, and analysis syntax are available on our GitHub <https://github.com/AGEexeter/paper-brain_iron_causes>.

### Supplementary Figures

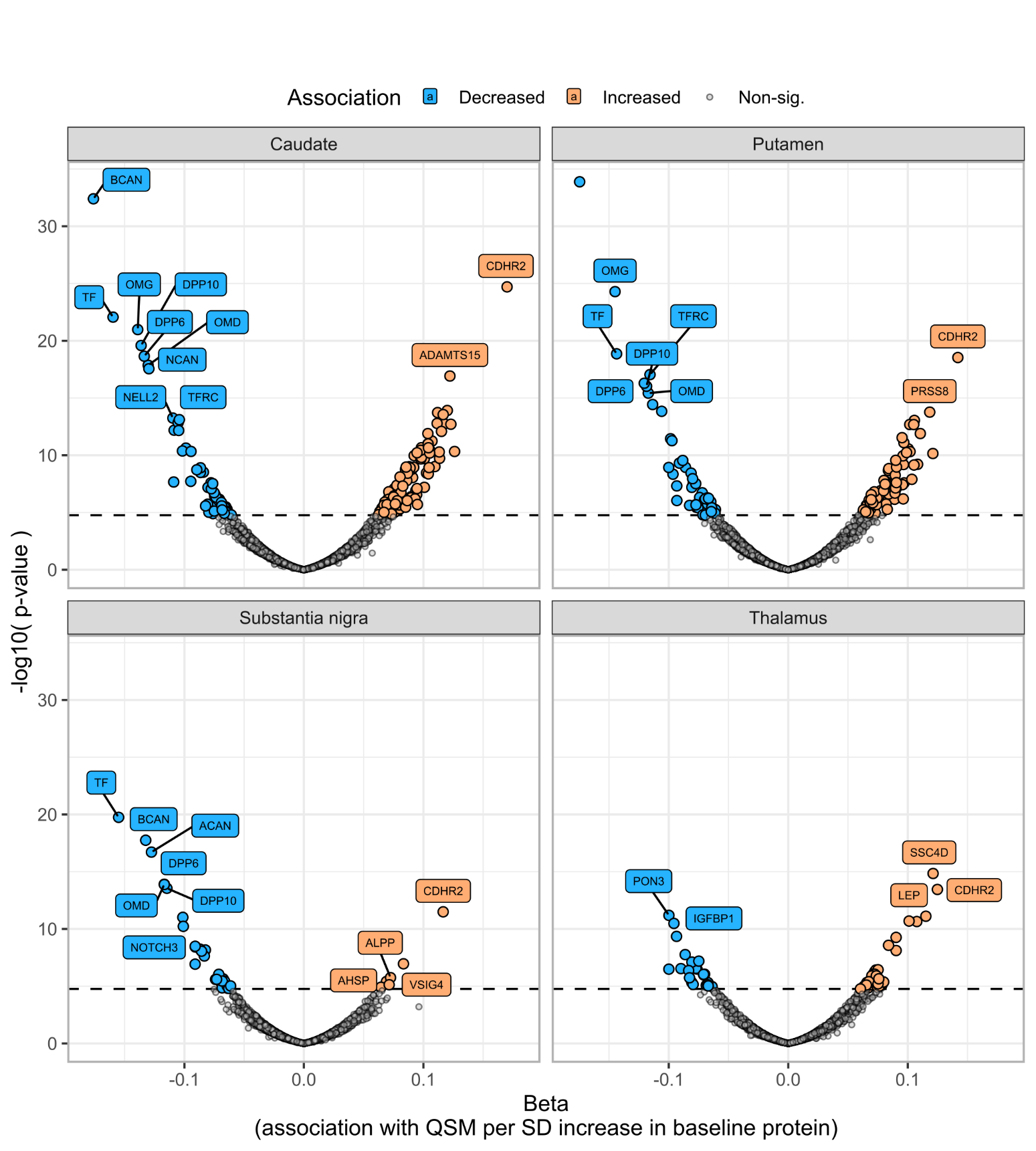

#### Figure S1. Baseline proteomic associations with subcortical QSM

Results from linear regression models in up to 3,614 UK Biobank participants, adjusted for age at baseline assessment, age at MRI assessment, sex, baseline assessment centre, MRI assessment centre, and whether the participant was UKB-PPP consortium selected. The beta is the standardized coefficient, representing the standard deviation (SD) difference in QSM at MRI assessment per standard deviation increase in baseline proteomic measure. The horizontal line indicates significant after Bonferroni correction for multiple statistical testing. See ST5 for details.

#### Figure S2. Prevalent disease associations with additional adjustments

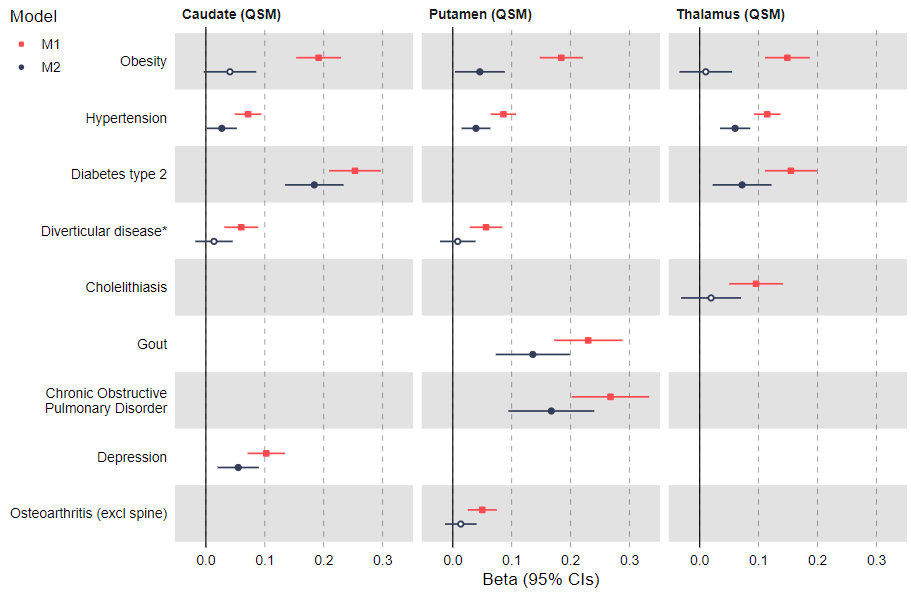

Model 1 = age, sex, assessment centre

Model 2 = M1 + ethnic background + education (highest qualification) + smoking status + alcohol (days per week) + physical activity (days per week with moderate activity) + meat consumption (days per week with red or processed meat intake) + waist circumference + systolic blood pressure

Results are only shown for disease-QSM pairs where there was a significant difference between the estimates from M1 and M2 (Fisher’s Z).

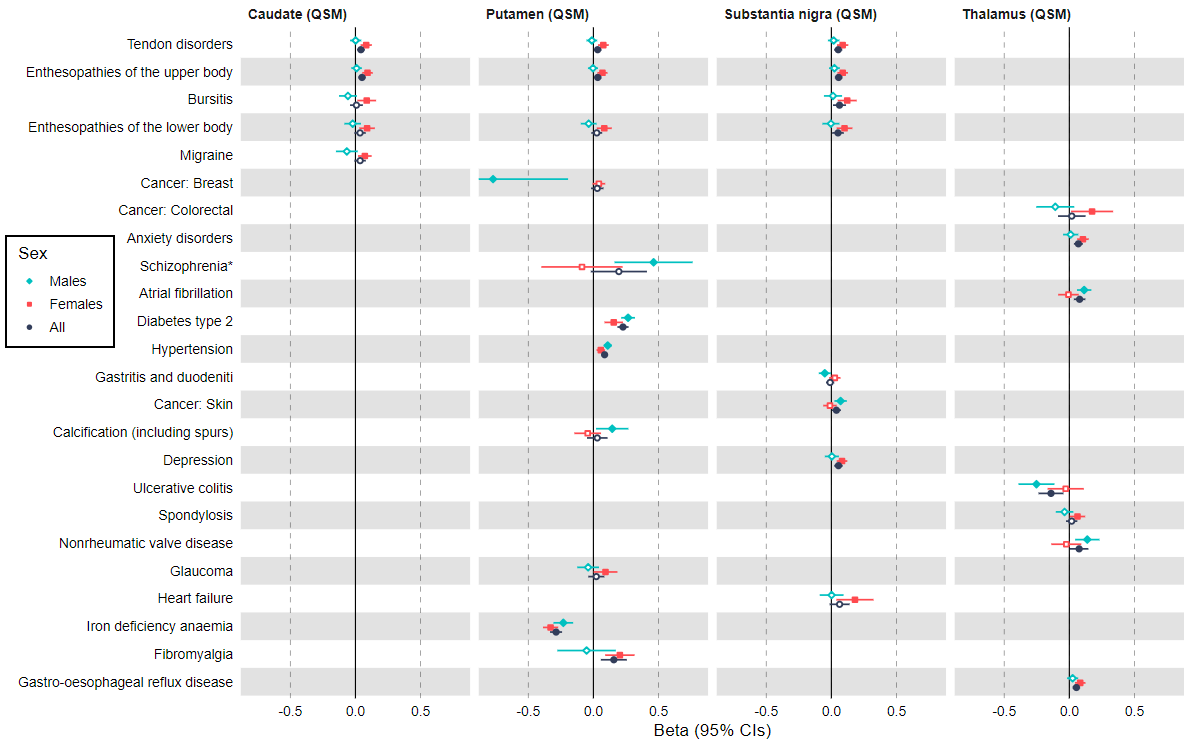

#### Figure S3. Prevalent disease associations stratified by sex

Results are only shown for disease-QSM pairs where there was a significant difference between the estimates in models stratified by sex (Fisher’s Z).

#### Figure S4. Scatter plot of SNP effect on calcium and SNP effect on caudate QSM

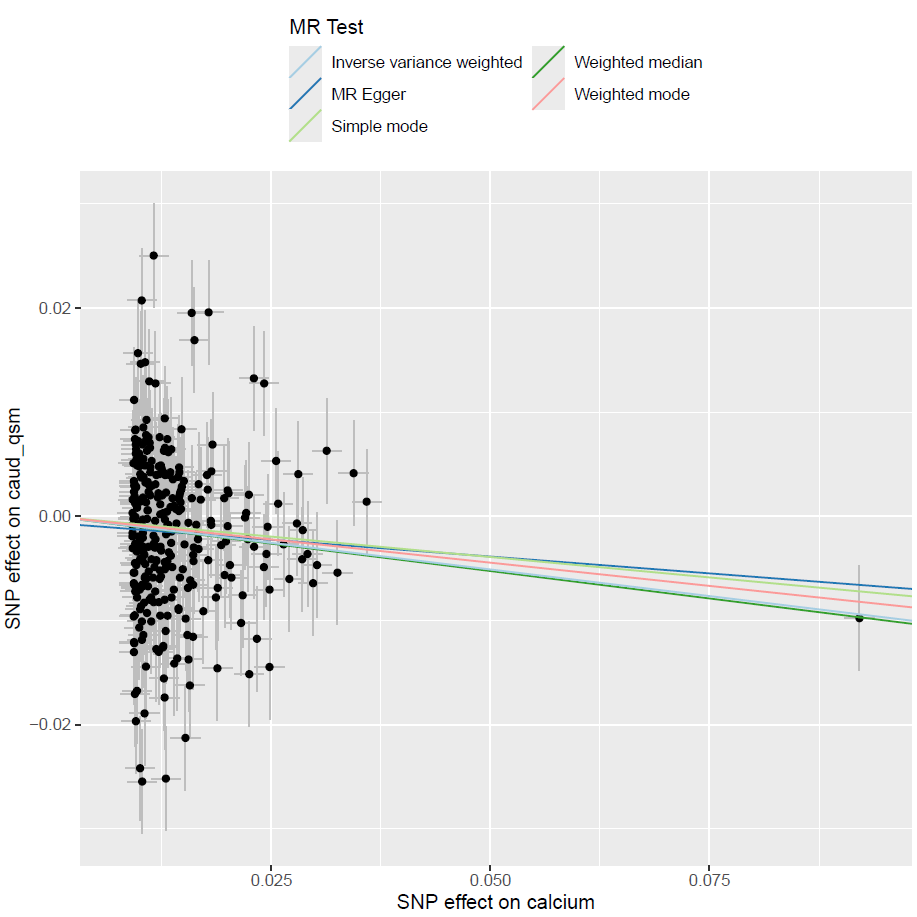

#### Figure S5. Scatter plot of SNP effect on alkaline phosphatase and SNP effect on substantia nigra QSM

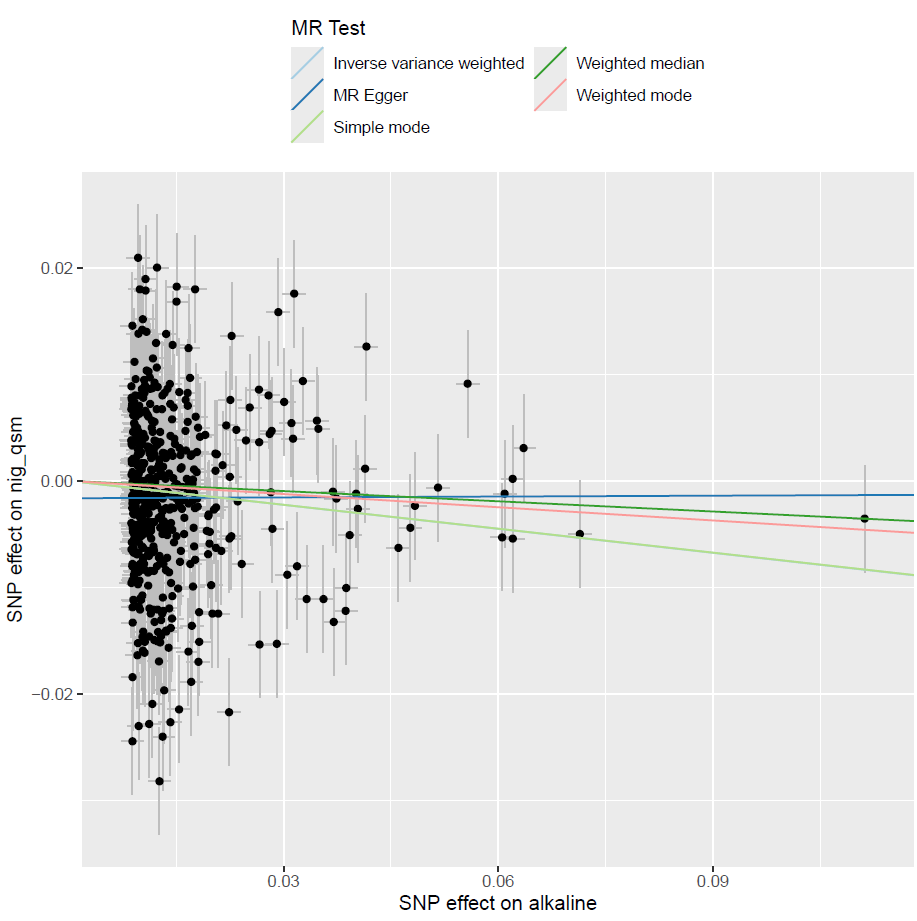

#### Figure S6. Scatter plot of SNP effect on urate and SNP effect on caudate QSM

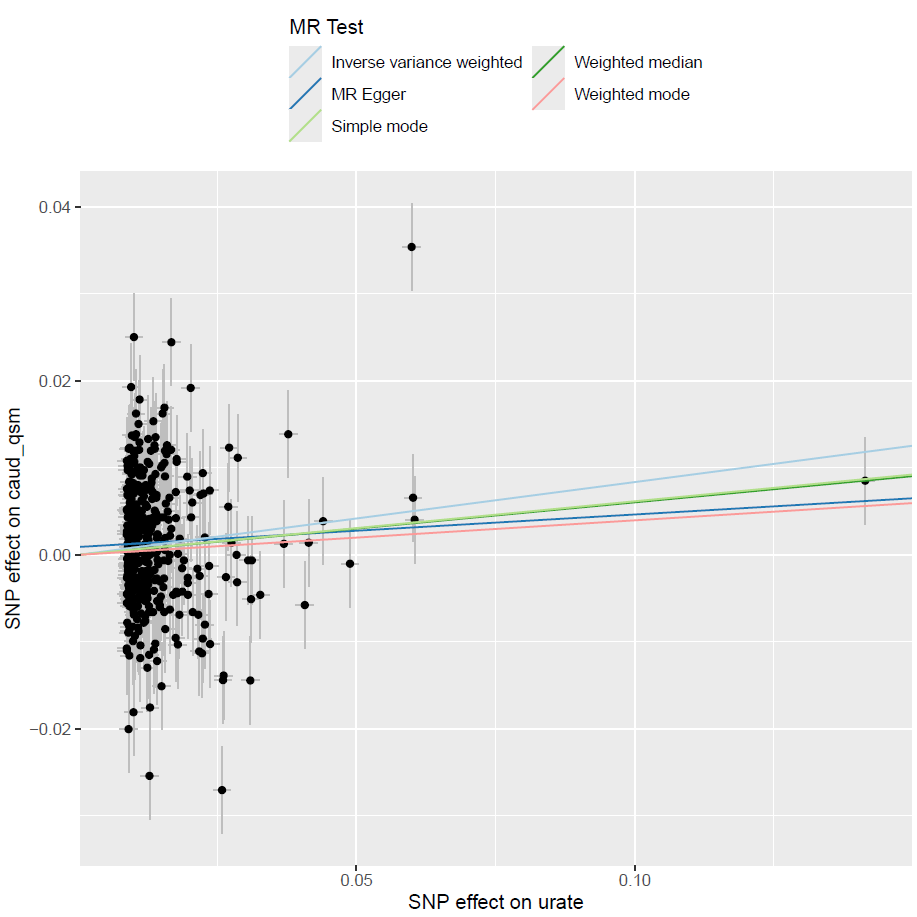

#### Figure S7. Scatter plot of SNP effect on urate and SNP effect on putamen QSM

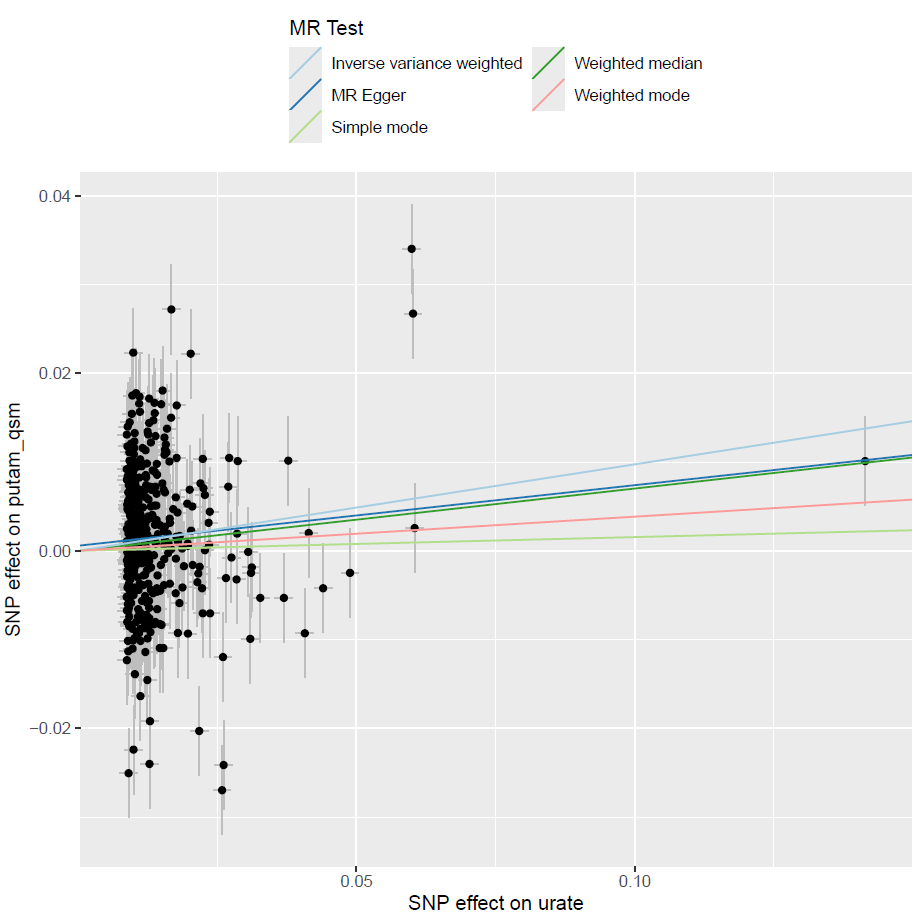

#### Figure S8. Scatter plot of SNP effect on asthma and SNP effect on caudate QSM

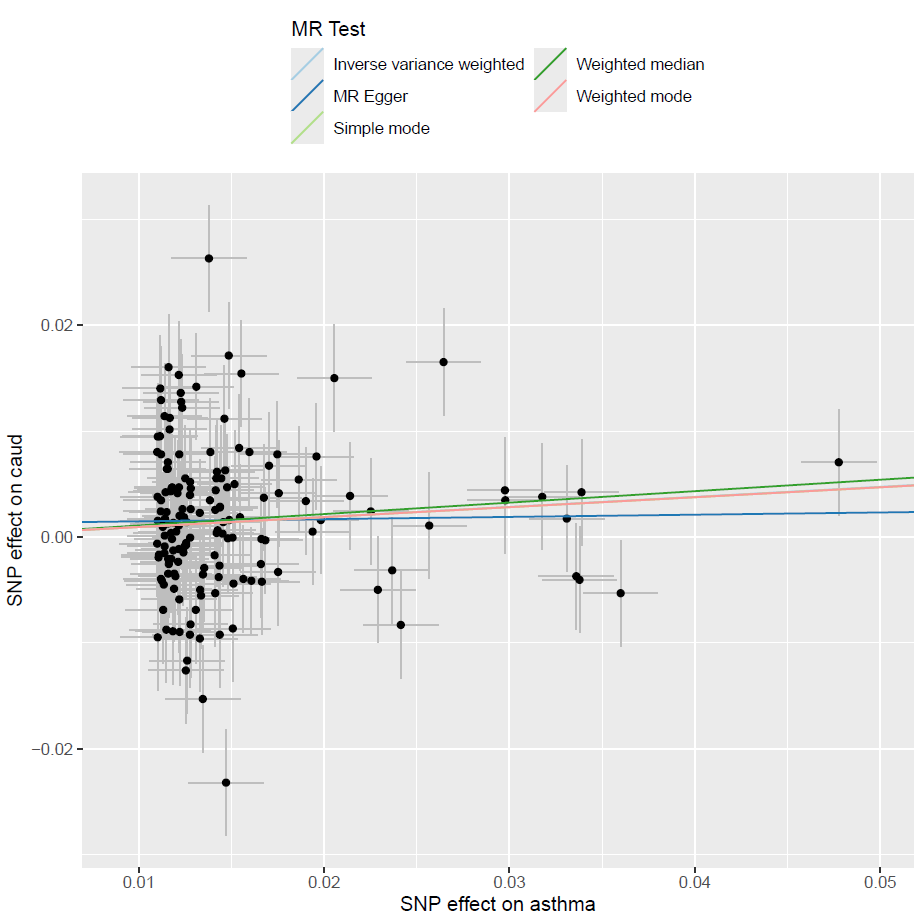

#### Figure S9. Scatter plot of SNP effect on asthma and SNP effect on putamen QSM

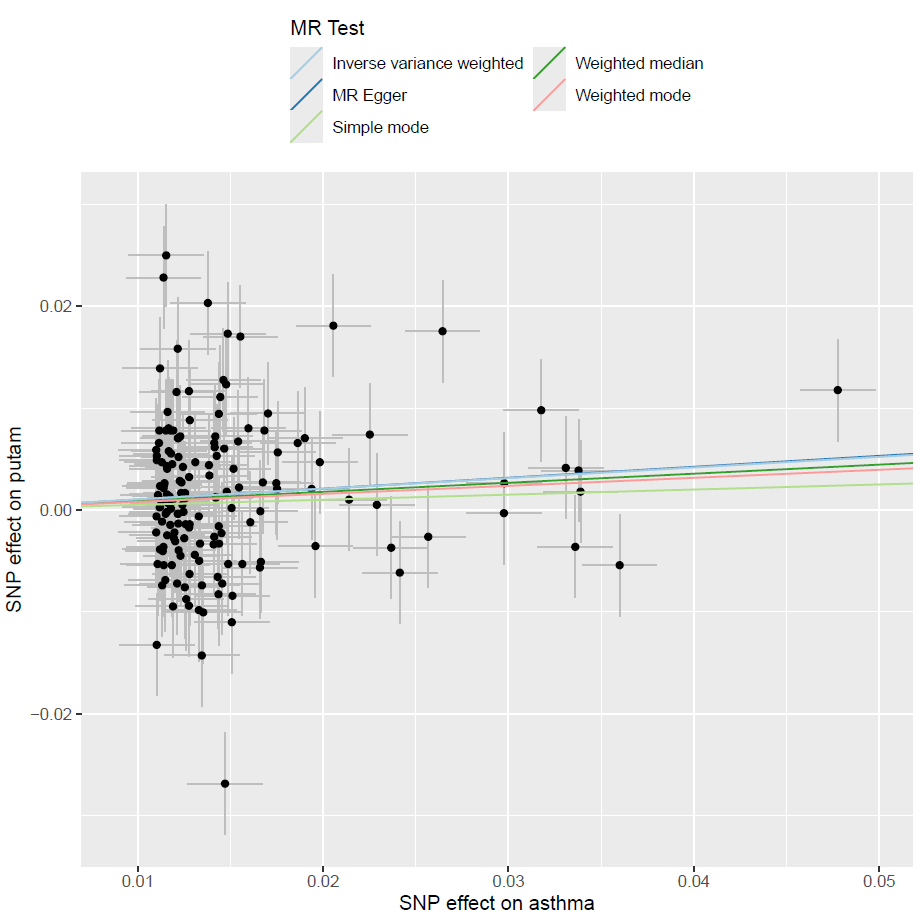

#### Figure S10. Scatter plot of SNP effect on COPD and SNP effect on putamen QSM

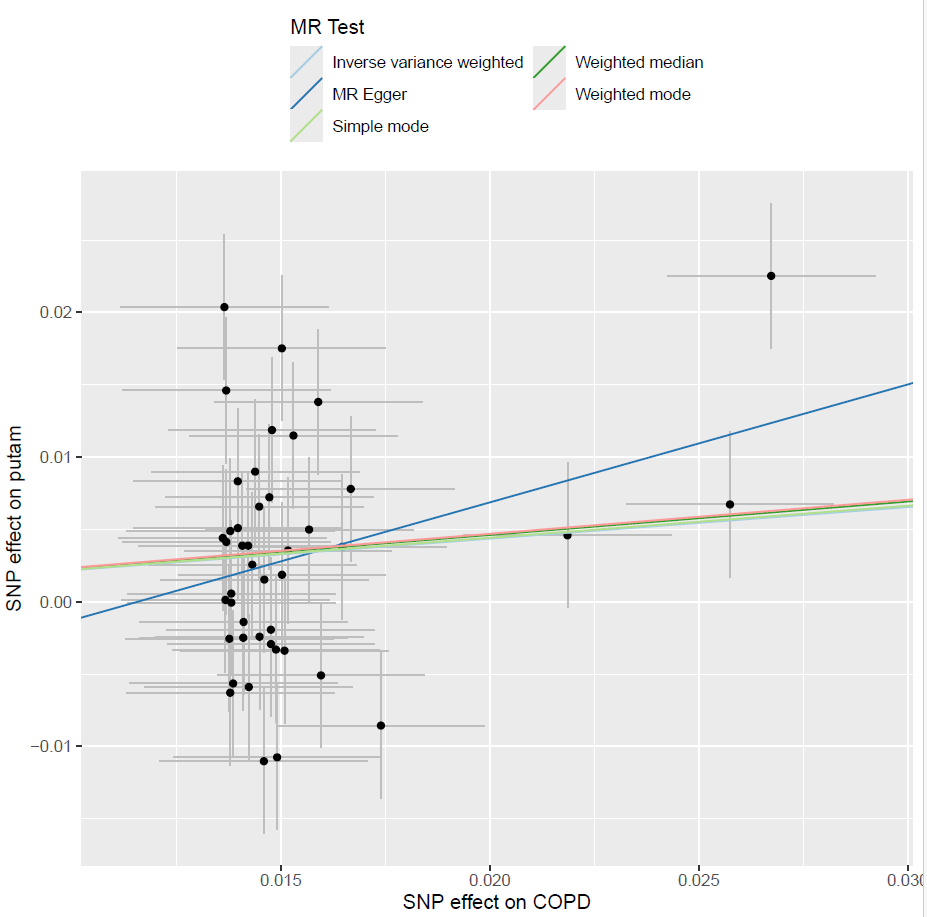

#### Figure S11. Scatter plot of SNP effect on type 2 diabetes and SNP effect on caudate QSM

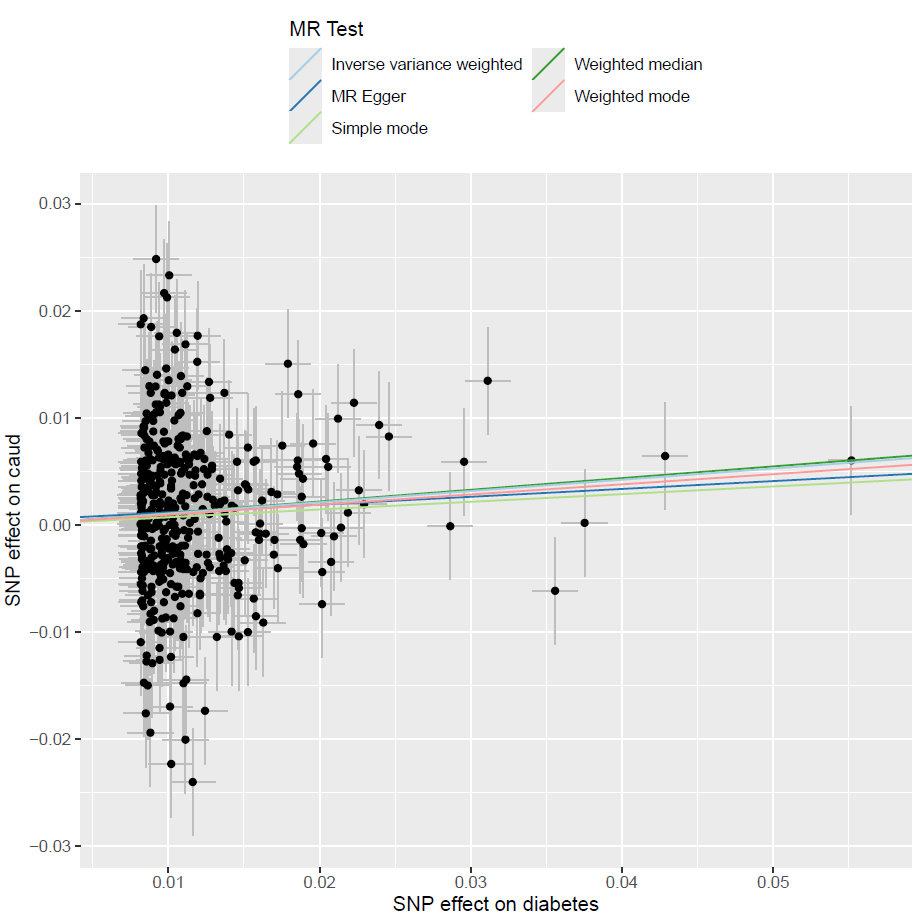

#### Figure S12. Scatter plot of SNP effect on type 2 diabetes and SNP effect on putamen QSM

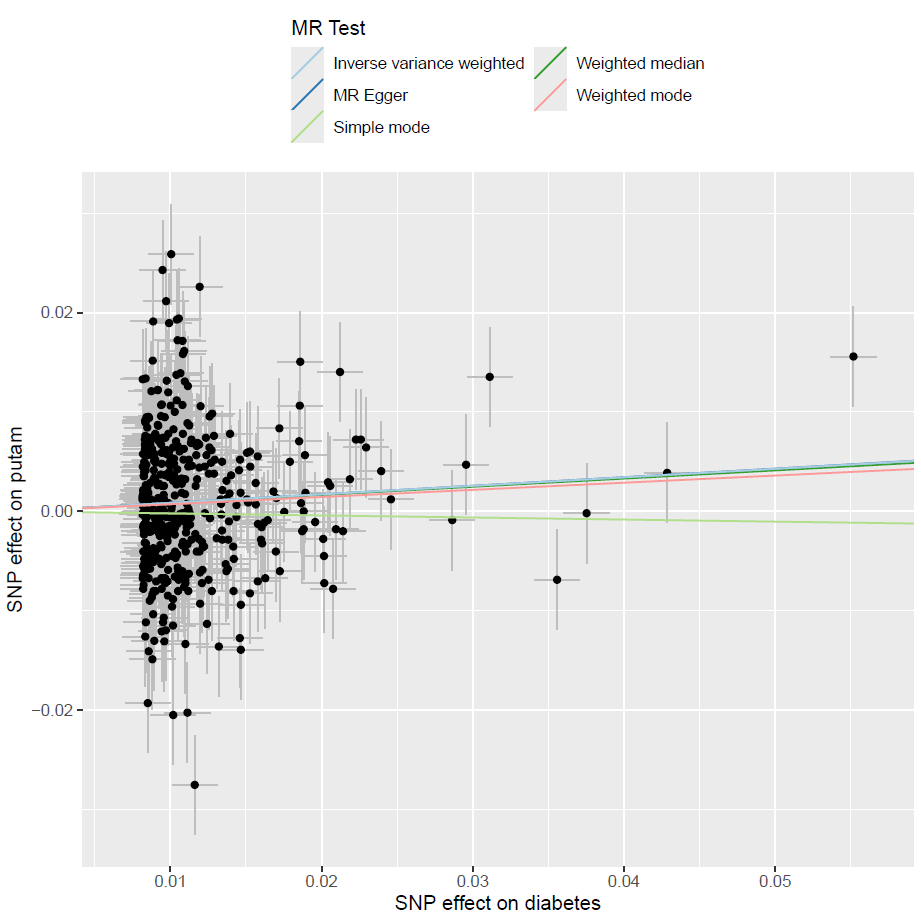
